# Prediction of Extubation Readiness Using Transthoracic Ultrasound in Preterm Infants

**DOI:** 10.1101/2020.04.26.20080739

**Authors:** Reem M. Soliman, Yasser Elsayed, Reem N. Said, Abdulaziz M. Abdulbaqi, Rania H. Hashem, Hany Aly

## Abstract

**Objective:** To test the hypothesis that a lung ultrasound severity score (LUSsc) and assessment of left ventricular eccentricity index of the interventricular septum (LVEI) by focused heart ultrasound can predict extubation success in mechanically ventilated preterm infants with respiratory distress syndrome (RDS).

**Design:** Prospective observational study of premature infants <34 weeks’ of gestation age supported with mechanical ventilation due to RDS. LUSsc and LVEI were performed on postnatal days 3 and 7 by an investigator who was masked to infants’ ventilator parameters and clinical conditions. RDS was classified based on LUSsc into mild (score 0–9) and moderate-severe (score 10–18). A receiver operator curve was constructed to assess the ability to predict extubation success. Pearson’s correlation was performed between LVEI and pulmonary artery pressure (PAP).

**Setting:** Level III neonatal intensive care unit, Cairo, Egypt.

**Results:** A total of 104 studies were performed to 66 infants; of them 39 had mild RDS (LUSsc 0–9) and 65 had moderate-severe RDS (score ≥10). LUSsc predicted extubation success with a sensitivity and a specificity of 91% and 69%; the positive and negative predictive values were 61% and 94%, respectively. Area under the curve (AUC) was 0.83 (CI: 0.75-0.91). LVEI did not differ between infants that succeeded and failed extubation. However, it correlated with pulmonary artery pressure during both systole (r=0.62) and diastole (r=0.53) and with hemodynamically significant patent ductus arteriosus (r=0.27 and r=0.46, respectively).

**Conclusion:** LUSsc predicts extubation success in preterm infants with RDS whereas LVEI correlates with high PAP.

## BACKGROUND

Respiratory distress syndrome (RDS) is the most common cause of neonatal morbidity and mortality in preterm infants.[1] The incidence of RDS is inversely proportional to gestational age, occurring in > 90% in preterm infants born < 28 weeks.[22] These infants are often supported with mechanical ventilation and rescued with surfactant administration via an endotracheal tube. Upon improvement in the course of RDS, infants are attempted for extubation to receive non-invasive mode of respiratory support such as continuous positive airway pressure (CPAP). Since there is no clear criteria to guide clinicians when to extubate, infants may not necessarily succeed the extubation attempt and are subsequently re-intubated. The exposure of infants to multiple intubations is not a safe practice. Studies demonstrated significant hemodynamic derangements that occur during intubation of premature infants.[3] Consequently, intubation has been associated with increased risk for intraventricular hemorrhage in premature infants.[4] Therefore, there is an unmet need to device an indicator for readiness to extubate in order to avoid the risks associated with re-intubation.

Lung ultrasound (LUS) can recognize a normal aerated lung in contrast to interstitial or alveolar patterns. In the last decade, lung ultrasounds have been increasingly used in critically ill patients, and evidence based international guidelines are published for the use of lung ultrasounds in adult critical care.[5] It is simple and raises no threat of radiation. However, the data on its feasibility and accuracy in the neonatal population are scarce.[6]

Echocardiography (ECHO) is considered the gold standard tool to detect anatomical cardiovascular defects, assess cardiac function, evaluate abnormal pulmonary circulation and estimate the response to therapeutic interventions. However, it requires specific skills and detailed training for a caregiver to perform neonatal ECHO.[7] Focused heart ultrasound is a simplified protocol of bedside ultrasound screening of pulmonary hypertension by measuring eccentricity index (EI). Left ventricle EI is a quantifiable measure of the amount of distortion of ventricular septal geometry that is related to increased right ventricular systolic or diastolic pressures and volumes. Left ventricular EI has been associated with pulmonary hypertension in children and adults but has not been validated in premature infants.[8]

This prospective study was conducted on premature infants diagnosed with RDS and supported with mechanical ventilation. Infants had point of care LUS and left ventricular EI measurements. The aim of this study was to test the hypothesis that a LUS severity score combined with left ventricular EI would predict success of extubation in mechanically ventilated preterm infants.

## DESIGN

### Patients

This prospective cohort study was conducted at the neonatal intensive care unit (NICU) of Kasr El Aini Hospital, a referral tertiary care university hospital. The study was approved by the Institutional Research Ethics Board and informed parental consents were obtained for all subjects before recruitment. Infants were included in the study if they fulfilled the following criteria: a) gestational age <34 weeks, b) postnatal age <48 hours, and c) supported with invasive mechanical ventilation for RDS. Infants were excluded if they had any of the following: a) major congenital anomalies, b) perinatal asphyxia, or c) hemodynamic instability managed by inotropic administration at the time of screening.

Demographic and clinical data were collected from the chart. Those data included the use of antenatal steroids, infant’s sex, gestational age (GA), birth weight (BW), small for gestational age status, Apgar scores at 5 and 10 minutes, perinatal confirmed infections, and intraventricular hemorrhage. Clinical parameters of hemodynamic stability including systolic and diastolic blood pressures, heart rate, urine output, and lactic acid at the time of assessment were collected.

LUS and transthoracic focused heart ultrasound (FHUS) were performed on postnatal days 3 and 7 for all recruited infants. The radiologist who performed the studies (R.H.) was masked to the clinical details of infants; likewise, the managing team in the NICU was blinded to the ultrasound results.

### Clinical respiratory parameters

Respiratory data were collected for all infants during the first 7 days of life. Data included mean airway pressure (MAP), fraction of inspired oxygen (FiO_2_), arterial blood gas measurements, total mechanical ventilation days, total oxygen days, and success/failure of extubation events. Oxygen saturation index (OSI) was calculated on postnatal days 3 and 7; OSI = MAP × FiO_2_ ×100 / arterial oxygen saturation (SpO_2_).[9] We stratified respiratory severity into mild, moderate, and severe based on OSI of ≤ 2.9, 3.0-6.4, and ≥6.5, respectively.[9]

### Extubation criteria

Patients were weaned to extubation according to standardized guidelines. Gradual weaning of FIO_2_ and peek inspiratory pressure (PIP) or tidal volume (VT) were performed guided by assessment of chest excursion, oxygen saturation and blood gases results. Infants were extubated when ventilatory rate was <25 breaths/min and PIP at 16-18 cmH_2_o that delivered desired VT. Infants were extubated to nasal CPAP or nasal non-invasive ventilation based on the observed work of breathing and presence of irregular breathing or apnea.

### Lung ultrasound protocol:[10]

A total of six lung zones were scanned, 3 on the right lung and 3 on the left lung. These zones are summarized in Figure 1. Four patterns of lung findings were described (Figure 2) with graded severity score; the lowest score is zero and the highest is 3 for each zone. Therefore, the total lung score is 18, which is the sum of the 3 lung zones on both right and left sides (3×3×2 = 18). The four patterns are identified as follows: Pattern 1: It shows transverse repetition of artifact (A-lines). This pattern reflects normal aerated lung. The lung in this pattern will have a few longitudinal artifact lines (B lines). Absence of B lines would suggest the diagnosis of pneumothorax. Pattern 2: It shows separated longitudinal artifact (B lines). The presence of more than 2 separated longitudinal B lines reaching the bottom of the screen would indicate the existence of interstitial edema. Pattern 3: It shows coalescent B lines that reflect alveolar edema. Pattern 4: It shows subpleural consolidation with air bronchograms. This pattern appears similar to pattern 3 but with white echogenic small dots of streaks representing cross-sections of small bronchi surrounded by fluid or collapsed alveoli.[8] In addition to these four patterns, the presence or absence of lung sliding is also noted during real-time US. Lung sliding indicates healthy movement between the lung tissue and the pleura. The absence of lung sliding indicates the presence of a pneumothorax.[10] Scoring of LUS are described in Figure 2 for each specific LUS pattern. Lung ultrasound severity score (LUSsc) graded from 0 (low risk) to 18 (high risk).[8]

**Figure 1:**
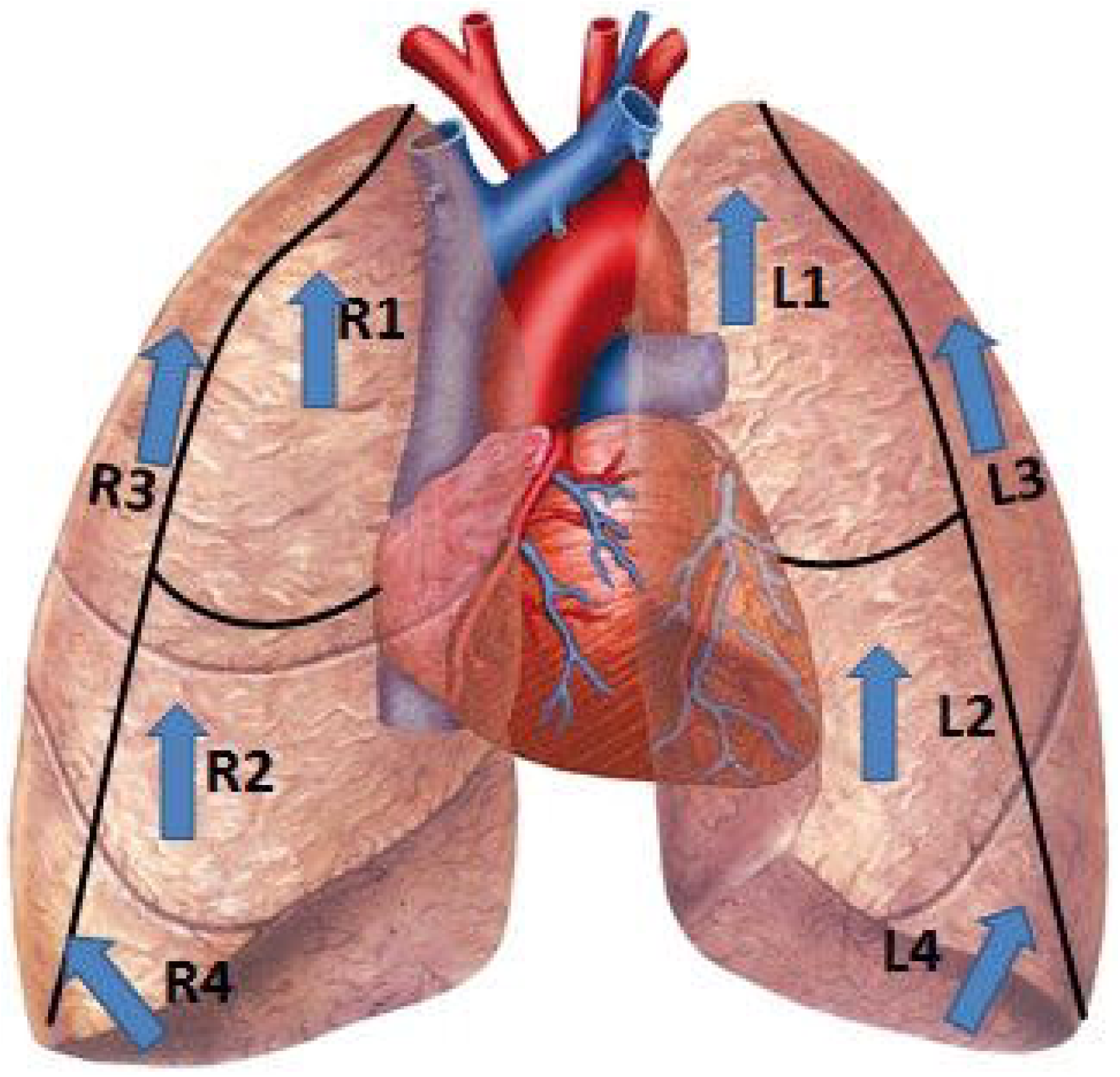
Lung zones used during ultrasound scanning R1: right upper anterior, R2: right lower anterior, R3: right lateral, R4: right costophrenic angle for assessment of pleural effusions, same for the left side. The direction of the arrows represents the direction of the ultrasound probe.

**Figure 2:**
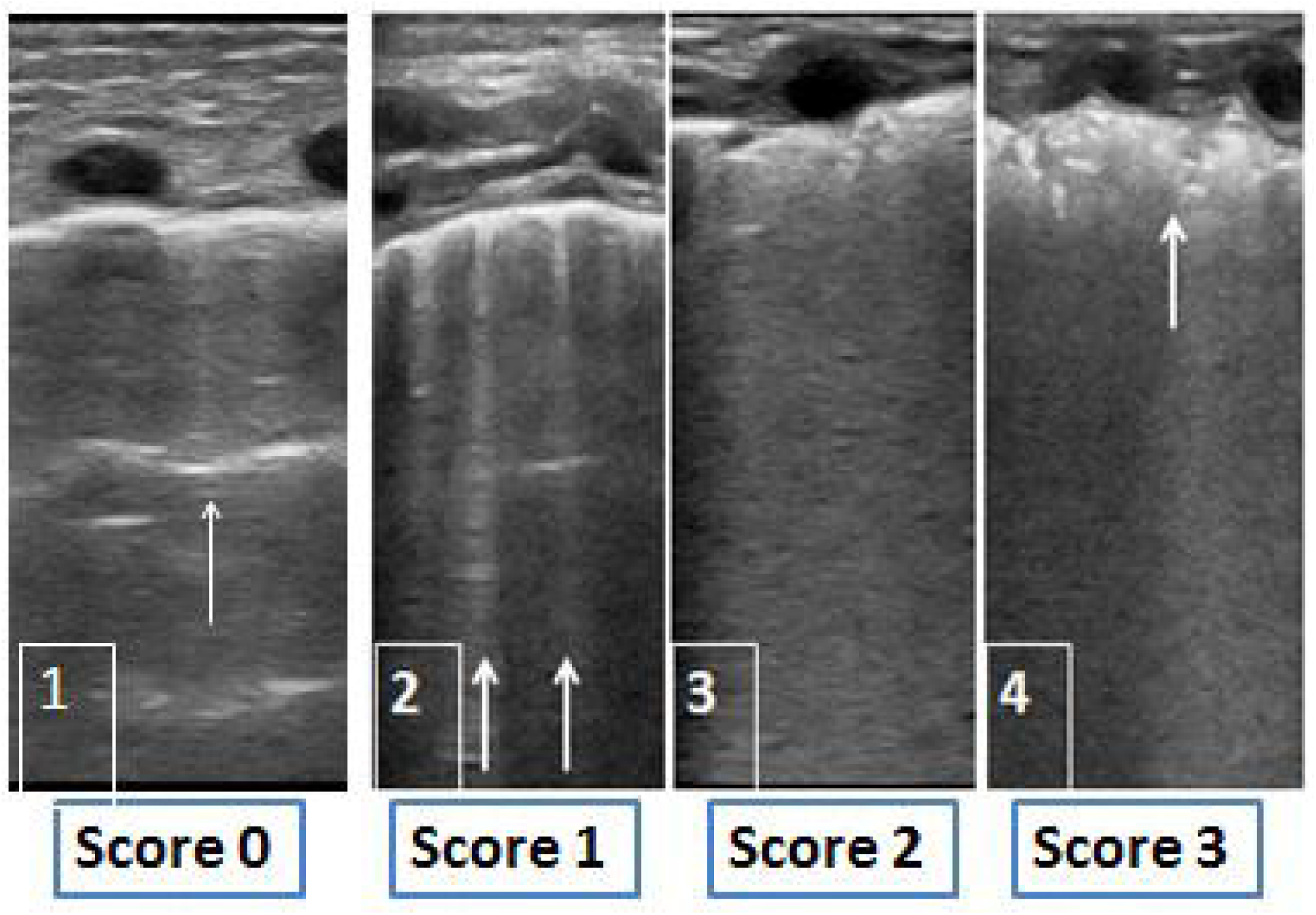
Four typical patterns of neonatal lung ultrasound 1) Normal lung areation with transverse repitation of pleural image which is A-line artifact (arrow), these patterns is given a severity score of zero. The absence of sliding in this study indicates the presence of a pneumothorax. 2) Longitudinal separated B lines (arrows) reaching the bottom of the image signifies interstitial fluid as in transient tachypnea of the newborn (TTN pattern). This pattern is given a severity score of 1. 3) Coalescent B lines with homogenous echographic view and thickened pleura as in respiratory distress syndrome (RDS pattern). This pattern is given a severity score of 2. 4) Subpleural air bronchogram (arrow) is seen in addition to the RDS pattern. This pattern us typically seen in infants with bronchopulmonary dysplasia (BPD pattern) and is given a severity score of 3 (the highest score).

### Focused heart ultrasound:[8]

FHUS was done on postnatal days 3 and 7 by the same radiologist (R.H.) who was experienced with neonates and was blinded to clinical details of the studied infants. Left Ventricular EI was measured at end-systole and end-diastole, from the parasternal short axis 2D image at the mid-papillary muscle level. The formula (EI = D2/ Dl) was used where: Dl is the ventricular diameter perpendicular to the interventricular septum bisecting D2, and D2 is the diameter parallel to the interventricular septum. EI was considered abnormal when the ratio was >1.0 and a higher EI was considered a marker of increased pulmonary vascular resistance. [11–13] In addition, full ECHO was done by pediatric cardiologist to screen for congenital heart disease and patent ductus arteriosus (PDA) and to measure PAP S assessment on day 3 of life.

In addition, full echocardiography was performed by a pediatric cardiologist that paralleled FHUS on postnatal day 3. This ECHO screened for congenital heart disease and patent ductus arteriosus (PDA), and measured pulmonary artery pressure. The two investigators who performed ECHO and FHUS did not communicate their evaluation and measurements.

### Statistical analysis

SPSS version 25 (SPSS Inc., Chicago, IL) was used to perform the statistical analysis. Data were presented as median with interquartile range or frequencies. Comparisons between groups were analyzed by Mann-Whitney U test. Frequencies were analyzed by chi-square test; p values < 0.05 were considered significant. Receiver operator curve (ROC) was constructed and predictive values were calculated. Pearson’s correlation was used to examine correlations of EI with PAP.

## RESULTS

A total of 104 point of care US were performed to 66 infants at a median (IQR) GA and BW of 30 (29–31) weeks and 1420 (1340–1880) g, respectively. Infants were classified into two groups based on LUS severity scores: Group-1 with LUS severity score (LUSsc) 0-9 which was considered as mild RDS and group-2 with LUSsc ≥ 10 which was considered as moderate to severe RDS. The two groups were comparable in demographic and clinical characteristics (Table 1).

**Table 1.** Demographic and clinical characteristics of the study population

|  | Group 1<br>LUS Score 0-9<br>n=(17) | Group 2<br>LUS Score 10-18<br>n=(49) |
| --- | --- | --- |
| Cesarean section <sup>a</sup> | 12 (70.6) | 32 (65.3) |
| Prenatal steroids <sup>a</sup> | 2 (11.8) | 3 (6.1) |
| Gestational age, week | 30 (29 - 31.5) | 30 (28.5 - 32) |
| Birth weight, kg | 1.45 (1.12 - 1.65) | 1.34 (1.12 - 1.66) |
| Male <sup>a</sup> | 10 (58.8) | 25 (51%) |
| Apgar at 1 min | 3 (1 - 5) | 3 (2 - 4) |
| Apgar at 5 min | 5 (4 - 7) | 5 (5 - 7) |
| Bronchopulmonary dysplasia <sup>a</sup> | 5 (29.4) | 7 (14.3) |

<sup>a</sup> Data are expressed in number (%), otherwise data are expressed in median (interquartile range).
There was no difference between the two groups in any of the variables ( $p>0.05$ ). LUS: lung ultrasound

Table 2 shows the respiratory characteristics of both groups; PIP was significantly higher in group-2 compared to group-1. Other ventilator parameters were comparable between the two groups.

**Table 2:** Respiratory characteristics of the two studied groups

|  | Group-1<br>LUS score 0-9 | Group-2<br>LUS score 10-18 | P value |
| --- | --- | --- | --- |

|  | N = (39) | N = (65) |  |
| --- | --- | --- | --- |
| Peak inspiratory pressure, cmH<sub>2</sub>O | 15 (14-16) | 16 (15-18) | 0.009 |
| Positive end expiratory pressure, cmH<sub>2</sub>O | 5 (5-6) | 5 (5-6) | >0.05 |
| Fraction of inspired oxygen, % | 35 (30-47.5) | 40 (35-55) | >0.05 |
| Failed extubation <sup>a</sup> | 15 (38.5) | 53 (81.5) | <0.001 |
| Oxygen saturation index | 2.14 (1.5-2.82) | 3.35 (2.7-5.375) | <0.001 |
| Duration of ventilation, day | 6 (5-8) | 8 (5.5-11) | 0.004 |
| Duration of oxygen support, day | 21 (8-33) | 14 (7-21) | 0.03 |

<sup>a</sup> Data are expressed in number (%), otherwise data are expressed in median (interquartile range).
LUS: lung ultrasound

LUSsc differed significantly between infants who succeeded extubation (n=36) and those who failed extubation (n=68): 7 (6–10) vs 12 (10.3 - 15), respectively; p<0.0001. OSI was also different between those who succeeded and failed extubation: 2.1 (1.5 - 2.6) vs 3.4 (2.7-4.7), respectively; p <0.0001. Duration of mechanical ventilation was significantly increased in infants who failed extubation: 5 (5 - 8) days vs 8.5 (8 - 9) days, p<0.0001.

LUSsc could predict extubation success at a cutoff point 11 with a sensitivity and a specificity of 91% and 69%; the positive and negative predictive values were 61% and 94%, respectively. Area under the curve (AUC) was 0.83 (CI: 0.75-0.91), Figure 3.

**Figure 3:**
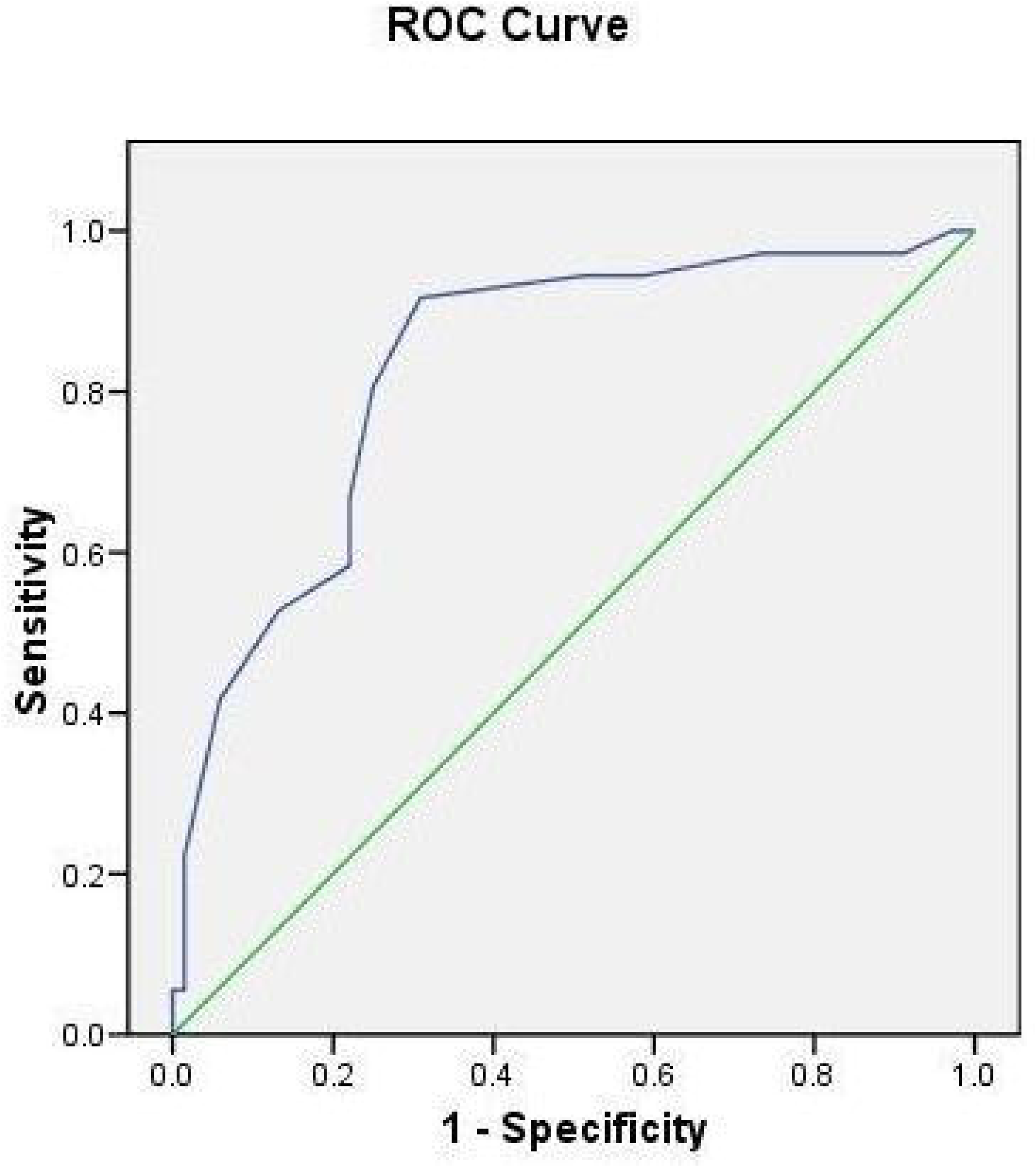
Lung ultrasound score for the prediction of extubation success Receiver operating characteristic (ROC) curve at a cutoff point 11 has a sensitivity and a specificity of 91% and 69%; the positive and negative predictive values are 61% and 94%, respectively. Area under the curve (AUC) is 0.83 (CI: 0.75-0.91)

Left ventricular EI did not differ between infants that succeeded and failed extubation. Its value during systole was 1.3 (1.2 – 1.58) vs 1.37 (1.2-1.5), p=0.94 and during diastole was 1.3 (1.13 – 1.48) vs 1.3 (1.2 – 1.56), p=0.75, respectively. LV EI correlated with PAP when measured during both systole and diastole (r=0.62, p<0.001 and r=0.53, p<0.001, respectively) and with hemodynamically significant PDA (r=0.27, p=0.03 and r=0.46, p<0.001, respectively). (Supplement Figures S1-S4)

## DISCUSSION

This study demonstrated the ability of point of care LUS to predict success of extubation in mechanically ventilated premature infants. Infants with higher LUSsc were supported with higher venitlatory pressure. Left ventiruclr EI did not differ among infants who succeeded or failed extubation. However, it correlated with PAP.

Infants with higher LUSsc were supported with significantly higher pressure of conventional mechanical ventilation. The use of higher PIP is indicative of decreased compliance in infants with severe RDS. A previous study of CPAP-supported infants demonstrated the need for higher pressure in infants with higher LUSsc.[10] Oxygen saturation index (OSI) was also increased in infants with severe RDS who had higher LUSsc. This finding agreed with previous studies that showed correlation of LUSsc with OSI and with SpO_2_/FiO_2_ ratio.[10, 13]

This study is the first to use LUSsc to predict success of extubation from mechanical ventilation in premature infants. Many lung ultrasound scores have been developed to assess lung aeration and guide respiratory care in adults especially those with restrictive lung disorders; LUS in this population is strongly recommended (level of evidence A).[14] Compared to adult literature, LUS has not been adequately addressed in neonates although it is easier owing to the small patients’ size and the absence of obesity or heavy musculature.[15] A few studies described the LUS patterns in common respiratory conditions in neonates, such as meconium aspiration syndrome, hyaline membrane disease, transient tachypnea of the neonate, and pneumothorax.[16–19] Other studies described the usefulness of LUS in predicting the need for invasive mechanical ventilation in infants supported with noninvasive ventilation,[20, 21] and the need for surfactant replacement in CPAP-supported extremely preterm neonates.[22] However, there is no study to predict success of extubation from mechanical ventilation. Giving the ease of its use at the bedside and the non-invasive nature of LUS, it is helpful to obtain LUSsc before extubation to avoid the exposure of infants to re-intubation should the extubation attempt fail. One of the main barriers to the more extensive use of the ultrasound technology in the NICU is the lack of efficient training solutions and the need to have structured quality-check assurance.[23] Once training is established the use of LUS can be the first-line imaging technique in preterm infants.[22]

Pulmonary hypertension in newborns (PHTN) is triggered by multiple etiologies including hypoxemia and underlying parenchymal lung diseases and can potentially hinder extubation.[24] Performing a full echocardiographic studies on all mechanically ventilated preterm infants would be exhaustive and may not consistently correlate with PHTN.[11] Left ventricle EI is a quantifiable measure of the amount of distortion of ventricular septal geometry due to elevated right ventricular systolic or diastolic pressures and/or volumes. Greater degrees of left ventricular EI have been associated with PHTN in children and adults, but have not been studied thoroughly in premature infants. The left ventricular EI has the added benefit of being easily measured from any short axis view of the mid-left ventricle.[11] Increased end-diastolic EI would indicate volume overload as in hemodynamically significant PDA and increased end-systolic EI may indicate right ventricular volume overload as in PHTN.[25] In the current study, systolic and diastolic EI correlated with PAP that was measured concomitantly by echocardiography. It correlated with the presence of PDA as well. However, end-systolic and end-diastolic EI did not correlate with OSI and did not prove to be valuable in predicting readiness for extubation.

This study has the strength of addressing the success of extubation trials that is a real challenge in premature infants. The study has some limitations including the lack of comparison of LUSsc with findings of MRI or CT scan of the chest. All LUS studies were performed with the same investigator; that is important for consistency and reliability of the score. However, the reproducibility of these findings by other scanners is required for generalization of these results. LUSsc addresses failure of extubation related to lung parenchymal disease. Other factors involved in extubation failure such as airway edema and apnea of prematurity can not be predicted with LUS.

## CONCLUSIONS

This study demonstrated the ability of LUS to predict success of extubation in premature infants with RDS with an AUC >80%. The use of this simple bedside non-invasive test can potentially avoid the exposure of premature infants to multiple extubation-reintubation cycles. This study provides preliminary data that can be used to design a prospective randomized trial aiming at comparing pulmonary outcomes of premature infants who are extubated with and without the assistance of LUS.

## Data Availability

Data is available upon request. Otherwise, it will not be publically available.

## CONFLICT OF INTEREST

All authors declare no financial or conflict of interest in relation to this research. Authors did not receive any funding for this research project.

**What is already known on this topic**

– Lung ultrasound is increasingly used in critically ill adults. It is simple and does not impose radiation. Its feasibility and accuracy in neonates are unknown.

**What this study adds**

– Lung ultrasound reliably predicts the success of extubation from mechanical ventilation in premature infants with RDS. Left ventricular eccentricity index correlates with pulmonary artery pressure.

